# Impact of COVID-19 and the postponement of the Tokyo 2020 Paralympic Games on Paralympic athletes and staff: A cross-sectional study

**DOI:** 10.1101/2022.12.14.22283406

**Authors:** Matthias Walter, Adam Mesa, Laura A. McCracken, Carolyn Barakso, Daniel Bueno Buker, Pablo Benavides González, Wanda Holtz, Conchita Jaeger, Tom E. Nightingale, Andrei V. Krassioukov

**Author notes:** **Correspondence to:** Dr. Matthias Walter, Dr. Tom E. Nightingale. These authors contributed equally to this work (i.e. shared first authorship) (Matthias Walter, Adam Mesa). These authors contributed equally to this work (i.e. shared senior authorship) (Tom E. Nightingale, Andrei V. Krassioukov).

## Abstract

We aimed to investigate the impact of the postponement of the Tokyo 2020 Paralympic Games on expected participants’ careers, COVID-19 history and mental health using an e-survey. Thirty-nine participants (median age 37 years; 16 females) from five countries responded between July 20^th^ and September 28^th^, 2021, of which 37 completed the survey, including 20 athletes and 11 coaches. All but two participants planned to attend the rescheduled Games in 2021 (95%). Ninety percent (35/39) had previously tested at least once for COVID-19, with six testing positive. While three had no symptoms, all six were moderately impacted. Scores (median; lower and upper quartiles; questionnaire) for depression (2; 0.75-4; PHQ-9) and anxiety (2; 0-5.25; GAD-7) were low. Scores for post-traumatic stress disorder (PTSD) (3.5; 1-11; IES-R) were also low, but four participants reported high scores indicative of clinical concern for PTSD. There was low emotional distress caused by postponement of the Games (2; 1-4.5), and moderately low fear of catching COVID-19 (3; 2-5.5) on 10-point (0 = none, 10 = extreme) rating scales. While overall this population appears relatively resilient, the postponement of the Games came at a cost for some athletes and coaches, specifically with regards to experiencing symptoms of PTSD.

## Introduction

The postponement of the Tokyo 2020 Paralympic games due to the COVID-19 pandemic was clearly a necessary safeguard measure to protect the health of athletes, professionals and public who were scheduled to compete in, work or to be a spectator at the Games. There are, however, potential negative consequences of this decision on the mental health and athletic or professional careers of these individuals. The postponement of the Tokyo 2020 Paralympic games is one component of the multifactorial impact of the COVID-19 pandemic on athletes and professionals, in addition to other factors such as reduced access to training and fitness centres, fear of infection and anxiety surrounding ability to recover if infected, and isolation from teams, communities and social support.^1^ The negative consequences of the COVID-19 pandemic and the postponement of the Tokyo 2020 Paralympic games are likely particularly salient for Paralympic athletes. Individuals with disabilities are disproportionately affected by government restrictions, more likely to experience worse outcomes from COVID-19,^2,3^ and are at higher risk for mood disorders.^4^ There is a need to investigate the effects of the COVID-19 pandemic and related postponement of the Tokyo 2020 Paralympic games on the athletes and professionals involved. Thus, we aim to investigate the impact of the COVID-19 pandemic and the postponement of the Tokyo 2020 Paralympic games on associated athletes, coaches, medical professionals, and members of National Paralympic Committees (NPCs).

## Methods

### Ethics

This study protocol was designed by an international team of clinicians, scientist, and coaches and was approved by the University of British Columbia Clinical Research Ethics Board (H20-04011).

### Study participants

A web-based survey was distributed to several NPCs who invited potentially eligible participants. Inclusion criteria: Athletes, coaches, physicians, medical professionals, and members of NPCs with an age of at least 18 years, who would have participated in Tokyo 2020 Paralympic Games (had they not been postponed). Exclusion criteria: those with a history of any cognitive disorders and/or traumatic brain injury that could impact the participants’ ability to follow instructions and answer questionnaires, and those who were not scheduled to participate in or support the Tokyo 2020 Paralympic Games.

### Survey

This survey included the following five components: 1) demographics, 2) personal COVID-19 history, 3) participation in Paralympic games (Tokyo 2020 and previous ones), 4) impact of postponement of Tokyo 2020 Paralympic Games on athletic performance or professional career, and 5) validated questionnaires including the patient health questionnaire (PHQ-9), generalized anxiety disorder assessment (GAD-7), and the impact of events scale-revised (IES-R). The IES-R, which ask participants about an individual event, was written to ask specifically of feelings around the postponement of the Tokyo 2020 Paralympic Games. The survey was offered to participants in five languages: English, French, German, Portuguese and Spanish. The survey was split into multiple screens based on the aforementioned components and to facilitate conditionally shown questions. Question and answer order was not randomized. All participants were provided with an informed consent form and had the opportunity to consent online. Participants were informed that the survey was expected to take approximately 30-45 minutes to complete, with a progress tracker at the top and a back button.

### Security

The survey was hosted on the platform ‘Qualtrics’, which complies with the BC Freedom of Information and Protection of Privacy Act (FIPPA) as the survey data is encrypted, kept secure and is stored and in Canada. This platform is the survey tool endorsed and used by our home institution. The security and privacy policy for ‘Qualtrics’ can be found at: https://it.ubc.ca/services/teaching-learning-tools/survey-tool/ubc-survey-tool-terms-use.

### Statistical analysis

Demographic information, history of COVID-19, participation in the Paralympic Games and the impact of the postponement of Tokyo 2020 Paralympic Games on participants’ athletic or professional careers will be presented with descriptive statistics. Data are presented as raw values. Results are presented as sum or median with 25% and 75% quartiles and range (i.e., min - max). Statistical analyses were conducted using R statistical software version 4.0.5 for macOS.

## Results

### Participants

A total of 39 participants (16 females [41%]; median age 39 years, 29 – 46, range 18 – 76) responded to our invitation between July 20^th^ and September 28^th^, 2021. Thirty-seven (95%) participants completed the whole online survey, while one provided answers in part (skipping the validated scales at the end of the survey). The majority of participants were Paralympic athletes (51%, n = 20). Demographics and characteristics of participants are highlighted in Table 1. The impact of the Paralympic Games postponement and respondents’ self-reported mental health is highlighted in Figures 1 and 2, with the full breakdown in Table 2.

**Table 1.** Demographics and characteristics of participants.

| **Characteristics** | **n = 39** |
| --- | --- |
| **Age in years** |  |
| ≤ 20 | 4 |
| 21-30 | 7 |
| 31-40 | 12 |
| 41-50 | 7 |
| 51-60 | 6 |
| 61-70 | 1 |
| 71-80 | 2 |
| **Sex** |  |
| Female | 16 |
| Male | 23 |
| **Country representation** |  |
| Argentina | 23 |
| Switzerland | 10 |
| Canada | 3 |
| Chile | 1 |
| Brazil | 1 |
| Other (not disclosed) | 1 |
| **Participants role regarding the Tokyo 2020 Paralympic games** | **n = 39** |
| Athlete | 20 |
| Coach | 11 |
| Member of the national Paralympic committee | 2 |
| Assistant to the team | 2 |
| Team physician | 1 |
| Team physical therapist | 1 |
| Veterinarian | 1 |
| Support staff | 1 |
| **Sport (athletes and coaches only)** | **n = 31** |
| Para athletics | 12 |
| Wheelchair tennis | 4 |
| Para table tennis | 4 |
| Wheelchair rugby | 3 |
| Para cycling | 2 |
| Para shooting sports | 2 |
| Para badminton | 1 |
| Para Judo | 1 |
| Para rowing | 1 |
| Para swimming | 1 |
| **Previous participation at Paralympic games (athletes and coaches only)** | **n = 31** |
| Yes | 16 |
| No | 15 |
| **Impairment / diagnosis according to IPC (athletes only)** | **n = 20** |
| Impairment of muscle power | 10 |
| Hypertonia | 3 |
| Limb deficiency | 2 |
| Leg length deficiency | 2 |
| Unknown (not disclosed) | 2 |
| Vision impairment | 1 |

**Table 2.**
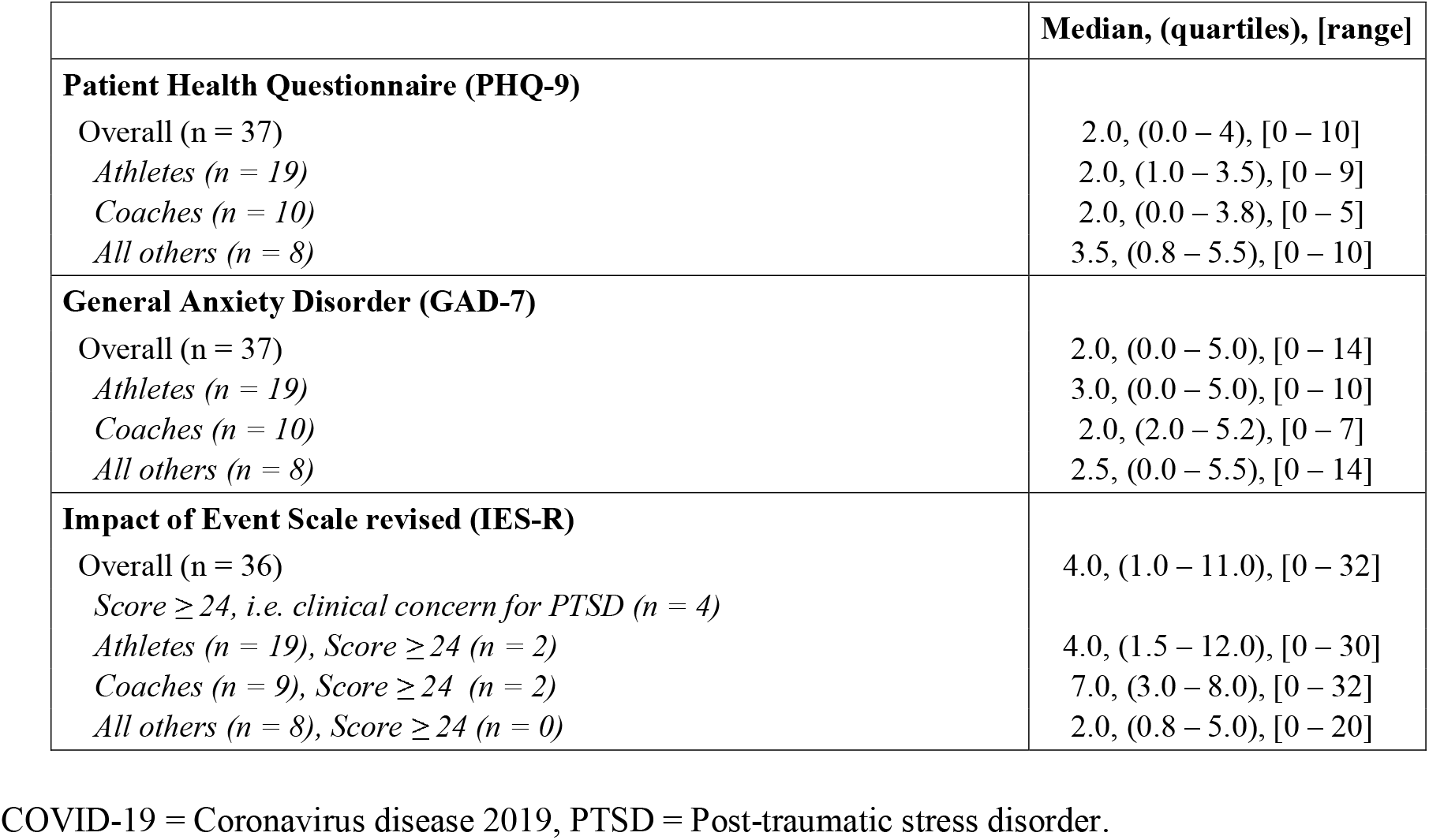
Overall impact of COVID-19 on Paralympians.

**Figure 1.**
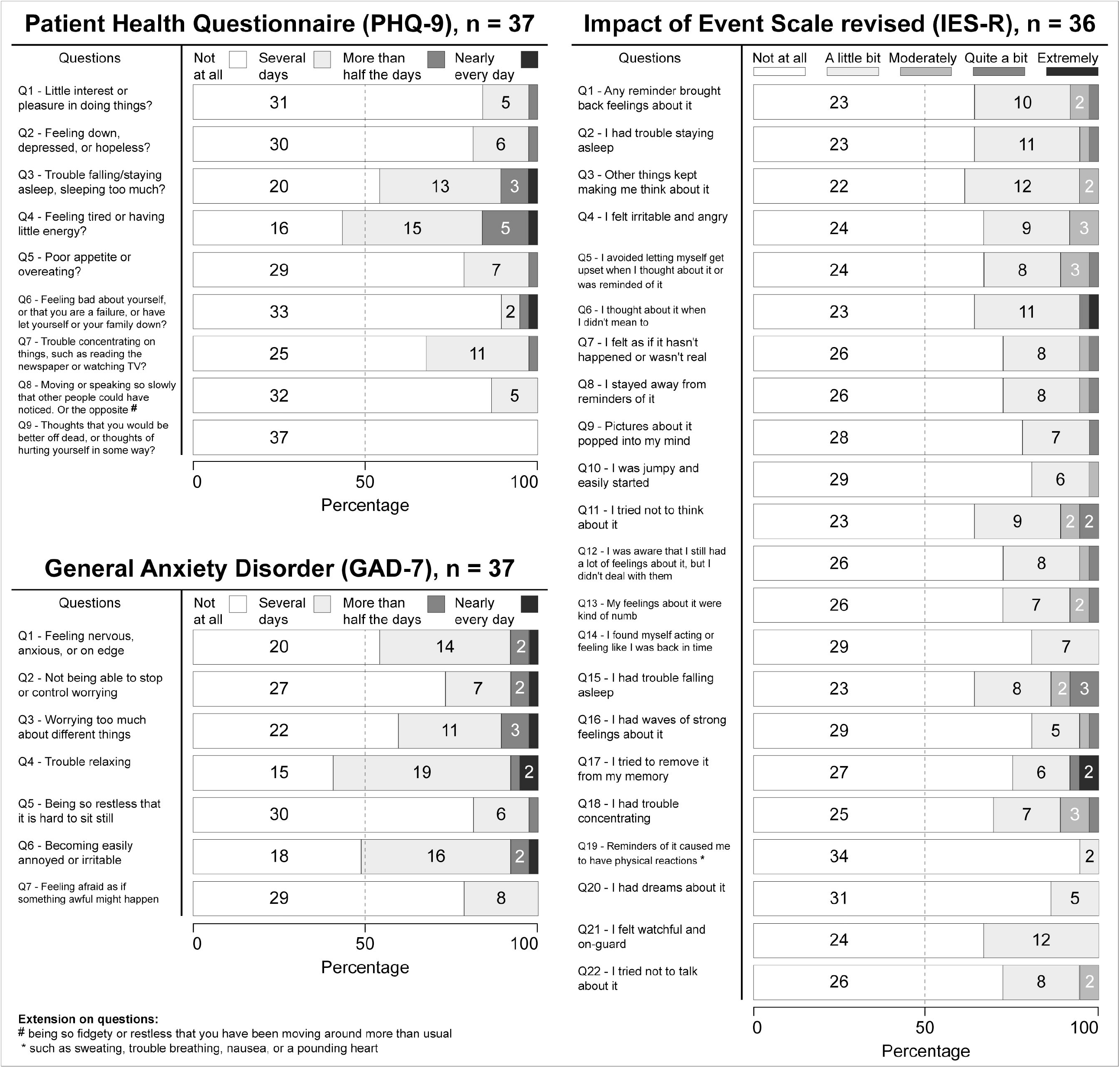
Distribution of Patient Health Questionnaire (PHQ-9), General Anxiety Disorder (GAD-7) and Impact of Event Scale revised (IES-R) responses. Q = question.

### COVID-19 infection and its impact on participants

Overall, 90% (35/39) of all participants were tested at least once for COVID-19. From that subgroup (n=35), only 6 (17%) tested positive (5 athletes and one coach from Argentina [=4], Brazil [=1], and Switzerland [=1]). Three participants did not experience any symptoms, two reported a mild illness (i.e., fatigue; fast and pounding heartbeat; sleep problems; shortness of breath, muscle/joint pain, memory/concentration problems, emotional instability), while one stated a moderate illness (i.e., some evidence of lower respiratory disease during clinical assessment or imaging; fatigue). Of these participants, the athletes were asked to rate “How much has your COVID-19 infection and post COVID-19 symptoms impacted the following area of your life?” from 0 - 2 (i.e., negligible impact), 3 - 7 (i.e., moderate negative impact) to 8 - 10 (i.e., significant negative impact). With regards to overall quality of life (n = 4), two reported a negligible impact, one a moderate negative impact, and one a significant negative impact. Inversely, with regard to their ability to continue training (n = 5), two reported a negligible impact, one a moderate negative impact, and two a significant negative impact. In addition, 97% (38/39) of all participants were immunized.

### Impact on mental health

As seen in Table 2, overall scores on the PHQ-9 and GAD-7 were low, with only one and two participants scoring moderately (≥10, i.e., probable depression or generalized anxiety disorder) on the PHQ-9 and GAD-7, respectively. Scores were similarly low on the IES-R, but 11% (4/36) scored highly (≥24, i.e., clinical concern for PTSD). Further, participants were asked to rate their fear of catching COVID-19 from 0 (no fear) to 10 (extreme fear), with responses being low overall (n = 37; 3; 2 – 5.5), including athletes (n = 19; 4; 3 – 7), coaches (n = 10; 3; 2 – 4), and all others (n = 8; 2; 1 – 3.5).

### Impact of the Tokyo 2020 Paralympic Games’ postponement on career

Given that all study participants would have participated in the Tokyo 2020 Paralympic Games had they happened, 95% (37/39) indicated they were also planning to participate in the 2021 Paralympic Games. The two participants not planning on participating in the 2021 Paralympic Games were a coach and medical professional. When asked how much activity levels (for athletes) or professional activities (for all others) had changed compared to before the pandemic started, 51% (19/37) participants reported slightly less (n = 15) or considerably less activity (n = 4) than before, while 24% (9/37) reported increases in activity. Of just athletes, a similar proportion reported slightly less (n = 9) or considerably less activity (n = 1). Additionally, 35% (13/37) of respondents felt the economic burden of the COVID-19 pandemic had affected their ability to train or perform professional duties, though there was no statistically significant correlation with the responses to the question on reduction in activity or work levels. At the time of survey completion, 81% of respondents (30/37) had resumed normal athletic training or normal professional activities. Of the seven who had not resumed normal activity, four reported more activity than before the pandemic, citing training facility closures (for one athlete and an assistant), discomfort in getting additional exposure to COVID-19 (for one athlete), and other tasks (for one coach). The reasons given by the other three who had not resumed normal activity levels and were doing less compared to prior to the pandemic were lockdowns (for one team assistant), “training pauses” (for one athlete), and loss of hospital resources (for one team physician).

## Discussion

The COVID-19 pandemic has caused global disruption, hardship, and challenges for many regardless of personal circumstance, and Paralympian athletes, coaches, and associated professionals are no exception. It has been reported that individuals with disabilities are more disproportionately affected by public health measures and are more vulnerable to worse outcomes if infected by COVID-19.^1,3^ Additionally, as elite athletes, Paralympians must balance strict training regimens, competitions, qualifiers and potentially even (future) career retirement.^5^ The postponement of the Tokyo 2020 Paralympic Games may have had a further negative impact. That said, no Paralympian works in isolation; there are coaches, team medical staff, assistants, NPC members, and other support staff who were potentially also disrupted. Thus, we set out to investigate this impact on participants’ athletic or professional career, personal COVID-19 history, and mental health. Recent studies have focused on athletes in the months immediately surrounding the postponement of the Olympic and Paralympic Games and what it meant for their mental health,^1,5,6^ but fewer have surveyed athletes around the time of the rescheduled Paralympic Games in 2021, like we did in this study.

While the postponement and the pandemic negatively affected participants, careers ultimately remained relatively unaffected, as all but two individuals indicated their participation in the rescheduled 2021 Paralympic Games. There may have been uncertainty around this for some in the initial weeks following the announcement.^5^ One recurrent challenge faced by athletes in other studies was the maintenance of physical activity or professional activities at pre-pandemic levels.^6,7^ This was due to early-pandemic public health measures such as facility closures,^6^ although this was less of a factor at the time of data collection in this report, as seen by a majority of respondents reporting pre-pandemic exercise levels.

Fortunately, the aforementioned public health measures were effective – COVID-19 infection rates were low among participants. In the few instances that it did occur, infection had a non-negligible impact on participants’ abilities to continue training, even for those who were asymptomatic. This was likely because of quarantine-minded public health measures (i.e., stay home when sick). However, these COVID-19-positive participants reported a low impact on their overall quality of life, and illness itself was mild or asymptomatic in all but one case.

Mental health issues reported among participants were also minimal overall, with low levels of probable depression and generalized anxiety disorder. This is a trend that has been seen in Paralympians relative to the general able-bodied population in other studies.^8^ Surprisingly, a tenth of athletes and coaches had PTSD-like symptoms towards the postponement of the games. While PTSD-like symptoms have been previously associated with pandemics^9,10^ and are highly prevalent in athletes from any source,^11^ our study data links the postponement of the Tokyo 2020 Paralympic Games to PTSD in only a relatively minor subsample of participants.

### Limitations

This was a convenience sample, which could have led to sampling and self-selection bias, with 64% of respondents coming from South America. However, there was representation from a range of other countries around the globe. Furthermore, responses were collected retrospectively, with responses collected over a year after the initial postponement of the Paralympics and at a point where participants were aware of their participation in the postponed games. Additionally, potential respondents who did decide not to pursue the rescheduled games may have been less likely to receive and participate in the online survey. This could partially explain the minimal effects reported in this cohort. Lastly, the study sample size was small and not all questions required participant responses, which explains the degree of missing data.

## Conclusions

Our findings reveal a population of resilient athletes, coaches, and associated professionals whose training and professional duties were at times disrupted by COVID-19 infection. This was demonstrated by the high number of participants who planned to or attended the rescheduled Paralympic Games in 2021, low infection rates and a relatively minor impact on quality of life for those infected with COVID-19. Participants mental health was predominantly good, with the exception of PTSD-like-symptoms in some of the cohort. This unexpected finding may prompt future studies and Paralympic organisations to implement strategies to support athletes and professionals, raise awareness of and minimize PTSD among their members, including around sporting event schedule changes.

## Data Availability

All data produced in the present work are contained in the manuscript.

## Acknowledgements

The authors would like to acknowledge all the participants of this study.

## Author contributions

All authors contributed to conception, study design, and development of surveys.

Authors 1 (MW) and 2 (AM) analysed the data.

All authors interpreted the data.

Authors 1 (MW) and 2 (AM) drafted the manuscript.

Authors 3 (LM), 4 (CB), 5 (DBB), 6 (PBG), 7 (WH), 8 (CJ), 9 (TEN), and 10 (AVK) revised the manuscript.

Authors 1 (MW) and 2 (AM) contributed equally (i.e. shared first author).

Authors 9 (TEN), and 10 (AVK) contributed equally (i.e. shared senior author).

